# Estimating pneumococcal carriage dynamics in adults living with HIV in a mature infant pneumococcal conjugate vaccine program in Malawi, a modelling study

**DOI:** 10.1101/2024.04.30.24306624

**Authors:** Joseph Phiri, Lusako Sibale, Lukerensia Mlongoti, Ndaona Mitole, Alice Kusakala, Mercy Khwiya, Thokozani Kayembe, Edwin Lisimba, Prosperina Kapwata, Ken Malisita, Chrispin Chaguza, Daniela M Ferreira, Deus Thindwa, Kondwani Jambo

**Author notes:** These authors contributed equally (Joseph Phiri, Lusako Sibale). These authors contributed equally (Deus Thindwa, Kondwani Jambo).

## Abstract

**Background:** Adults living with human immunodeficiency virus (ALWHIV) taking antiretrovirals (ART) have higher pneumococcal nasopharyngeal carriage and disease than adults without HIV (HIV-). To assess factors influencing high pneumococcal carriage prevalence and generate evidence base for evaluating future pneumococcal conjugate vaccine (PCV) strategies in ALWHIV, we estimated pneumococcal carriage acquisition and clearance rates in a high transmission and disease-burdened setting, at least 10 years after introducing infant PCV13 in routine immunisation.

**Methods:** We collected longitudinal nasopharyngeal swabs from age-and sex-matched 18–45-year-old HIV-adults, ALWHIV with ART experience of more than 1 year (ART>1y) or less than 3 months (ART<3m) from communities around Blantyre, Malawi. Samples were taken at baseline, and then weekly during the 16 visits over the study period. We employed classical culture microbiology to detect pneumococcal carriage and determined pneumococcal serotypes using latex agglutination. We fitted trajectories of serotype colonisation to multi-state Markov models to capture the dynamics of pneumococcal carriage adjusting for age, sex, number of household children under 5 years-old (<5y), social economic status (SES) and seasonality.

**Results:** At baseline, 65 adults were enrolled in each of the three HIV groups irrespective of pneumococcal carriage status, totalling 195 adults of whom 51.8% were females, 25.6% cohabited with >1 child <5y, and 41.6% lived in low SES. Median age was 33y (interquartile range [IQR]: 25-37y). Baseline pneumococcal carriage prevalence of all serotypes as 31.3% of which non-PCV13 serotypes (NVT) (26.2%) was higher than PCV13 serotypes (VT) (5.1%). In a multivariate longitudinal analysis, pneumococcal carriage acquisition was higher in females than males (NVT [Hazard Ratio [HR]: 1.53, 95%CI:1.17-2.01]; VT [1.96, 1.11-3.49]). It was also higher in low than high SES (NVT [1.38, 1.03-1.83]; VT [2.06, 1.13-3.77]), in adults living with 2+ than 1 child <5y (VT [1.78, 1.05-3.01]), and in ALWHIV on ART>1y than HIV-adults (NVT [1.43, 1.01-2.02]). Moreover, ALWHIV on ART>1y cleared pneumococci slower than HIV-adults ([0.65, 0.47-0.90]). Residual VT 19F and 3 were highly acquired although NVT remained dominant.

**Conclusions:** The disproportionately high point prevalence of pneumococcal carriage in ALWHIV on ART>1y is likely due to impaired nasopharyngeal clearance resulting in prolonged carriage. Our findings provide baseline estimates for comparison of pneumococcal carriage dynamics after new PCV strategies in ALWHIV are implemented.

**Author summary:** We assessed rates of pneumococcal serotype carriage acquisition and clearance by fitting multi-state Markov models to pneumococcal colonisation trajectories comprising 3,152 nasopharyngeal samples from 195 adults aged 18-45 years in Blantyre, Malawi. Substantial acquisitions of VT and NVT in females and those living under low socioeconomic status were estimated, in addition to VT acquisition among adults living with at least two children in the house and NVT acquisition among ALWHIV on ART>1y. ALWHIV on ART>1y cleared overall carriage, and NVT in particular, slower than their HIV-counterparts. Residual VT serotypes 19F and 3 were highly acquired whereas 19A, 3, and 6A were carried for longer durations, still, NVT serotypes remained dominant, suggesting that PCV strategy in ALWHIV should consider expanded serotype coverage to tackle the remaining preventable burden of pneumococcal carriage and subsequent disease. The contribution of NVT carriage to the disproportionately high carriage prevalence in ALWHIV is substantial, though the underlying causal drivers for prolonged duration of carriage in ALWHIV on ART>1y warrant further investigation. We generate the evidence base for evaluating future pneumococcal vaccine strategies in ALWHIV.

## Introduction

Streptococcus pneumoniae (pneumococcus) commonly colonises the nasopharynx (NP) of children and adults [1]. Pneumococcal colonisation precedes disease, such as pneumonia, meningitis, and bacteraemia [1–3], and is also prerequisite for transmission [1]. The pneumococcus causes excessively high pneumococcal carriage and disease burden in adults living with human immunodeficiency virus (ALWHIV) on antiretroviral therapy (ART) compared to adults without HIV (HIV-) [4, 5], despite the substantial coverage of ART and suppression of viral load [6, 7]. Paradoxically, the higher pneumococcal carriage prevalence among ALWHIV with longer than shorter ART experience remains unexplained in this setting [8].

Pneumococcal conjugate vaccines (PCVs) protect against carriage due to specific vaccine-targeted (VT) serotypes, thereby interrupting VT transmission and reducing VT disease risk [9, 10]. In November 2011, Malawi introduced the 13-valency infant PCV (PCV13) into the national extended programme on immunisation (EPI) using a three-primary dose schedule without a booster (3+0; one dose at 6, 10, and 14 weeks of age) [11]. Despite at least 12 years of more than 90% infant PCV13 coverage among age-eligible children [12], in the absence of a direct PCV vaccination program for ALWHIV [13], there is evidence of residual VT-carriage prevalence and VT-invasive pneumococcal disease (VT-IPD) incidence in children and ALWHIV in Malawi [14–17].

A change of infant PCV schedule from 3+0 to 2+1 (one primary dose at 6, 10, and booster dose at 36 weeks of age) or 2+1+1 (2+1 and one additional booster dose at 60 weeks of age) to enhance herd immunity, or direct routine PCV vaccination of ALWHIV has been suggested as potential vaccine strategies to eliminate persistent VT pneumococcal carriage risk and VT disease in ALWHIV [18]. Thus, to better assess the impact of a new vaccination strategy against VT carriage and disease among ALWHIV on ART, longitudinal studies are needed to generate the evidence base of pneumococcal serotype dynamics before the introduction of a new vaccine strategy [2, 19]. This may improve our understanding of serotypes that are frequently acquired or prolongedly carried, determinants of VT and NVT carriage acquisition and clearance, post-PCV serotype replacement, and the choice of vaccine product with the greatest potential to further reduce pneumococcal carriage and subsequent disease.

Here, we leveraged data from a longitudinal study of natural pneumococcal colonisation (Nasomune) among ALWHIV on ART and HIV-adults in Blantyre, Malawi, to estimate pneumococcal carriage parameters to inform transmission dynamic models of alternative vaccine strategies in ALWHIV. In particular, we estimated pneumococcal serotype-specific and vaccine-serotype group acquisition and clearance rates, as well as associated factors among ALWHIV using multi-state Markov transition models.

## Methods

### Ethics approval

Nasomune study nasopharyngeal (NP) samples were obtained from each Malawian adult through written consent. Study ethics approval was granted by the Malawi National Health Sciences Research Ethics Committee (NHSRC) (21/24/2680) and the Liverpool School of Tropical Medicine Research Ethics (21-035).

### Data description

Nasomune study data were collected between 17 September 2021 and 11 December 2023 in Blantyre, Malawi. Using a random sampling approach, individuals were enrolled from different communities across Blantyre of whom 65 were HIV-, 65 ALWHIV with at most 3 months ART experience (ART<3m), and 65 ALWHIV with at least 1 year ART experience (ART>1y). HIV infection status was determined according to the double rapid test algorithm in Malawi [20]. Inclusion criteria included asymptomatic adults aged 18 to 45 years living with at least one child under 5 years old. Adults with 4 weeks prior use of antibiotics (except cotrimoxazole), history of smoking, pregnancy, and current respiratory track illness were excluded from the study [21].

NP swabs were taken at baseline (visit 1) and then weekly during the 16 visits of the study period, resulting in 3,152 total NP samples from 195 individuals. The swabs were tested for the presence of pneumococci using the World Health Organisation (WHO) NP sampling procedure and standard microbiological culture [22]. Serotyping of every positive pneumococcal sample was done using latex agglutination, based on picking a single colony, to identify serotypes or serogroups [23]. Pneumococcal carriage density was measured using microbiological culture serial dilutions207 (comparable to quantitative polymerase chain reaction [qPCR208]) on gentamicin-sheep blood sugar agar plate (5µL gentamicin/mL) and results reported as colony forming units per millilitre (CFU/ml) [21, 24, 25]. On enrolment and during follow-up, clinical and demographic characteristics of the study participants were recorded which included antibiotic use during follow-up, pneumococcal carriage density, age, sex, number of children under 5 years old (<5y) living in the house, socioeconomic status (SES) based on owning different functioning items [3], and third-level administrative unit location where the study participant resided.

### Continuous-time time-homogeneous multi-state Markov models

We adapted a previously published Markov modelling framework to fit three variants of continuous-time time-homogeneous muti-state Markov models to individual-level trajectories of pneumococcal colonisation during the study period, assuming a susceptible-infected-susceptible (SIS) model structure [26]. The first model estimated total carriage dynamics regardless of specific serotypes and vaccine serotype category, the second model was expanded to capture VT and NVT carriage dynamics separately, and the last model was further expanded to also assess individual serotype carriage dynamics. Since multiple serotype carriage was not tested in this study due to use of latex agglutination serotyping method, we assumed that at any time-point t, an individual can only carry a single dominant serotype and be in a colonized state carrying pneumococcus (model 1) or separately VT and NVT (model 2) or any individual serotype (model 3) abbreviated as *I_g_*, or be in a uncolonized state (*S*) where g is the subscript for a carriage state. Thus, the transition intensities between {*S* and *I_g_*} can be described by transition matrices Q_l_ for model 1 (g=2), Q_2_ for model 2 (g = 2,3), or Q_3_ for model 3 with 16 individual serotypes (g = 2,3,4, …, 17) (Fig S1). To ensure model convergence due to limited data points, model 3 was limited to capture 16 carriage states, each corresponding to serotype 15A/B/C/F, 7A/B/C, 3, 11A/B/C/D/F, 23A/B, 17A/F, 19F, 10A/B/C/F, 20, 6C, 19A, 9A/L/N, 6A, combined identified VT (1, 4, 9V, 14, 18C, 23F), combined identified NVT (22A/F, 33A/B/C/D/F, 18A/B/C/F, 12A/B/F, 19B/C, 8, and 6D) and unidentified NVT.

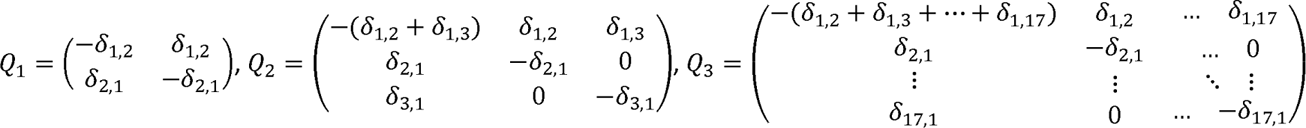

Our models describe acquisition and clearance rates of overall carriage, VT and NVT carriage or serotype-specific carriage from *S* to *I_g_* and from *I_g_* to *S*, captured by transition matrix entries *δ*_1,*g*_ and *δ*_1,*g*_, respectively. The effects (β) of a vector of clinical and demographic characteristics (*z_i_*) of *i*^th^ individual on acquisition and clearance rates are only estimated in model 1 and model 2 and not in model 3 due to limited data points. Thus, *β* is modelled using hazard rates [27] e.g., *δ_mn_*(*z_i_*(*t*))= *δ_mn_*^(0)^exp (β*_mn_^T^z_i_*((*t*)) over all transitions (*T*) where *m, n* = {1,2,3} refer to being in state n at time *t*>0 given that the previous state was *m*. Since acquisition of pneumococci is not observed for individuals already carrying pneumococci at baseline, we assume that their baseline rates of acquisition are similar to steady state rates over the study period. Our model also assumes that the future colonisation state is independent of its history beyond the current state [27]. We assume that the time spent in each state is exponentially distributed [26], thus pneumococcal carriage duration is estimated as the inverse of clearance, allowing precise estimation of pneumococcal carriage episode. To obtain acquisition probabilities, the matrix *P* is computed through matrix exponential, *P*(*t*) = exp (*Q*(*t*)), over a constant *Q* through the study period. To adjust for potential changes in pneumococcal carriage intensities over time due to seasonality, we include in model 1 and model 2 a binary term for hot-wet and cool-dry seasons representing a typical divide of Malawi’s climate over months of November-April and May-October[8], respectively (Fig S1).

### Likelihood estimation

To fit the Markov model, the likelihood is constructed as the product of probabilities of transition between observed states, over all individuals i and observation times j, assuming that sampling times are ignorable e.g., the fact that a particular observation is made at a certain time does not implicitly give information about the value of that observation.

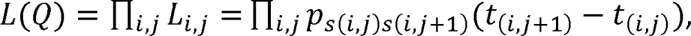

Where each *L_i,j_* is the entry of the transition probability matrix *P*(*t*) at *s*(*i, j*) row and *s*(*i, j* + 1) column evaluated at time *t*=*t* _(*i,j*+l)_ -*t* _(*i,j*)_. The likelihood *L*(*Q*) is maximised under a log scale to compute estimates of *δ_mn_* using Bound Optimisation By Quadratic Approximation (BOBYQA) algorithm implemented by msm R package [27, 28].

## Results

### Participant and sample description

At baseline, 65 individuals were enrolled in each group of HIV-, ALWHIV on ART<3m, and ALWHIV on ART>1y, totalling 195 participants of whom 5.1% and 26.2% carried VT and NVT, respectively. One hundred and one adults (51.8%) were females, 25.6% lived with at least two children <5y, and 41.6% were in low SES. The median participant age was 33 (interquartile range [IQR]: 25-37, range: 18-45), and pneumococcal carriage density was 10,720 CFU/ml (IQR: 1,005-82,075). We estimated carriage prevalence by diving the number of positive samples by the number of swabs taken per visit, HIV status and/or ART group. The baseline prevalence of pneumococcal NVT and VT carriage was generally higher for ALWHIV on ART>1y (33.8% and 7.7%) compared to ALWHIV on ART<3m (24.6% and 3.1%) or HIV-adults (20.0% and 4.6%). Likewise, the baseline median pneumococcal carriage density was higher for ALWHIV on ART>1y (13,400 CFU/ml; IQR: 520-67,838) compared to ALWHIV on ART<3m (9,548 CFU/ml; IQR 2,387-247,900) or HIV-adults (8,208 CFU/ml, IQR: 1,884-149,494) (Table 1).

**Table 1.**
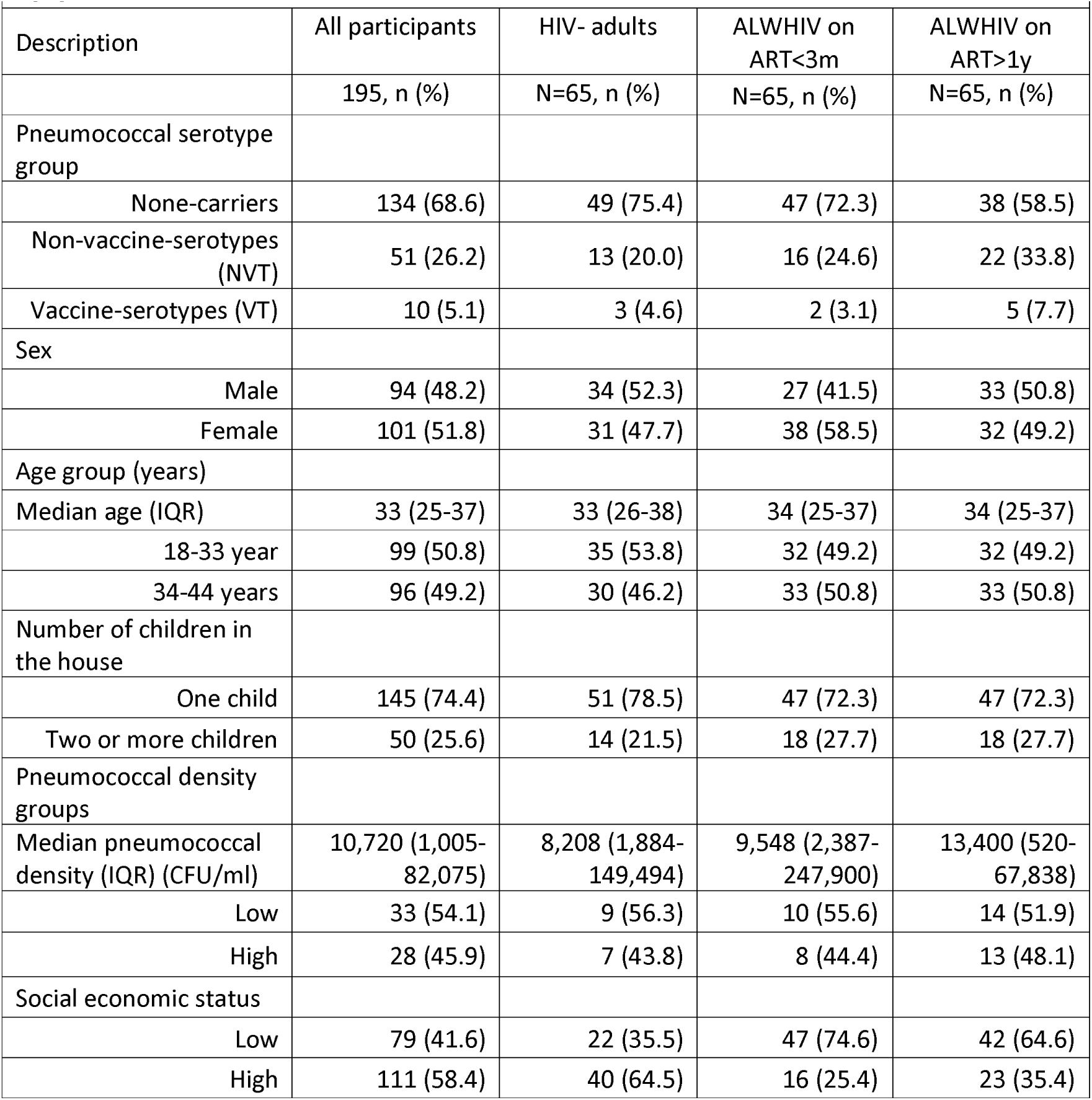
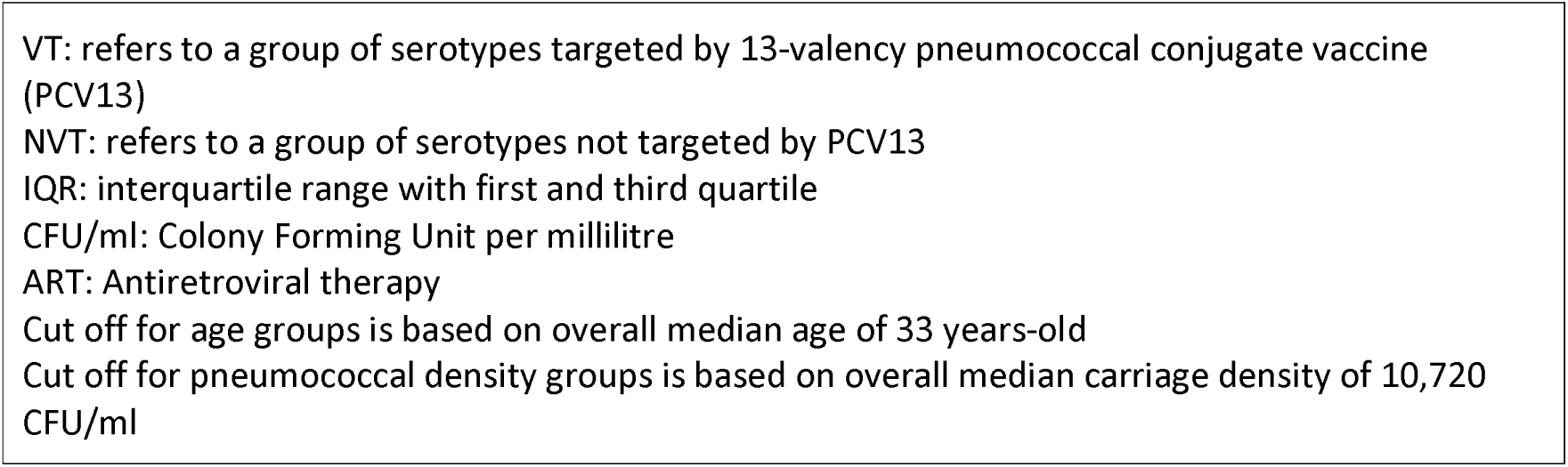
Baseline characteristics of adults without HIV (HIV-), adults living with HIV (ALWHIV) on ART<3 months (m), and ALWHIV on ART>1 year (y), who were recruited during in a pneumococcal nasopharyngeal swabbing study in Blantyre, Malawi between September 2021 and December 2023.

|  |  |  |  |  |
| --- | --- | --- | --- | --- |
| Description | All participants | HIV- adults | ALWHIV on ART<3m | ALWHIV on ART>1y |
|  | 195, n (%) | N=65, n (%) | N=65, n (%) | N=65, n (%) |
| Pneumococcal serotype group |  |  |  |  |
| None-carriers | 134 (68.6) | 49 (75.4) | 47 (72.3) | 38 (58.5) |
| Non-vaccine-serotypes (NVT) | 51 (26.2) | 13 (20.0) | 16 (24.6) | 22 (33.8) |
| Vaccine-serotypes (VT) | 10 (5.1) | 3 (4.6) | 2 (3.1) | 5 (7.7) |
| Sex |  |  |  |  |
| Male | 94 (48.2) | 34 (52.3) | 27 (41.5) | 33 (50.8) |
| Female | 101 (51.8) | 31 (47.7) | 38 (58.5) | 32 (49.2) |
| Age group (years) |  |  |  |  |
| Median age (IQR) | 33 (25-37) | 33 (26-38) | 34 (25-37) | 34 (25-37) |
| 18-33 year | 99 (50.8) | 35 (53.8) | 32 (49.2) | 32 (49.2) |
| 34-44 years | 96 (49.2) | 30 (46.2) | 33 (50.8) | 33 (50.8) |
| Number of children in the house |  |  |  |  |
| One child | 145 (74.4) | 51 (78.5) | 47 (72.3) | 47 (72.3) |
| Two or more children | 50 (25.6) | 14 (21.5) | 18 (27.7) | 18 (27.7) |
| Pneumococcal density groups |  |  |  |  |
| Median pneumococcal density (IQR) (CFU/ml) | 10,720 (1,005-82,075) | 8,208 (1,884-149,494) | 9,548 (2,387-247,900) | 13,400 (520-67,838) |
| Low | 33 (54.1) | 9 (56.3) | 10 (55.6) | 14 (51.9) |
| High | 28 (45.9) | 7 (43.8) | 8 (44.4) | 13 (48.1) |
| Social economic status |  |  |  |  |
| Low | 79 (41.6) | 22 (35.5) | 47 (74.6) | 42 (64.6) |
| High | 111 (58.4) | 40 (64.5) | 16 (25.4) | 23 (35.4) |

VT: refers to a group of serotypes targeted by 13-valency pneumococcal conjugate vaccine (PCV13) NVT: refers to a group of serotypes not targeted by PCV13 IQR: interquartile range with first and third quartile CFU/ml: Colony Forming Unit per millilitre ART: Antiretroviral therapy Cut off for age groups is based on overall median age of 33 years-old Cut off for pneumococcal density groups is based on overall median carriage density of 10,720 CFU/ml

During the follow-up visits, NVT carriage prevalence was mostly highest in ALWHIV on ART>1y (range: 7.2-13.2%) than in ALWHIV on ART<3m (5.9-11.6%) or HIV-adults (2.1-7.3%), whereas VT carriage prevalence remained similar in the three groups at 0.5-2.7%. For aggregated samples across all visits, NVT carriage prevalence remained higher in ALWHIV on ART>1y (10.6%, 95% Confidence Intervals [CI]: 9.6-11.8) than ALWHIV on ART<3m (9.1%, 95%CI: 8.2-10.2) or HIV-adults (5.8%, 95%CI: 5.0-6.7). Among NVT carriers, the median carriage density was higher among ALWHIV on ART>1y than HIV-adults. In contrast, the median carriage density was higher among ALWHIV on ART<3M than ALWHIV on ART>1y carrying either VT or NVT. In addition, NVT samples from ALWHIV on ART>1y dominated among those who lived with at least two children <5y (35.5%), who were 18-33y (33.3%), from low SES (38.7%) and did not use antibiotics (34.5%). Conversely, NVT samples from ALWHIV on ART<3m were highest among females (33.6%) (Fig 1).

**Figure 1.**
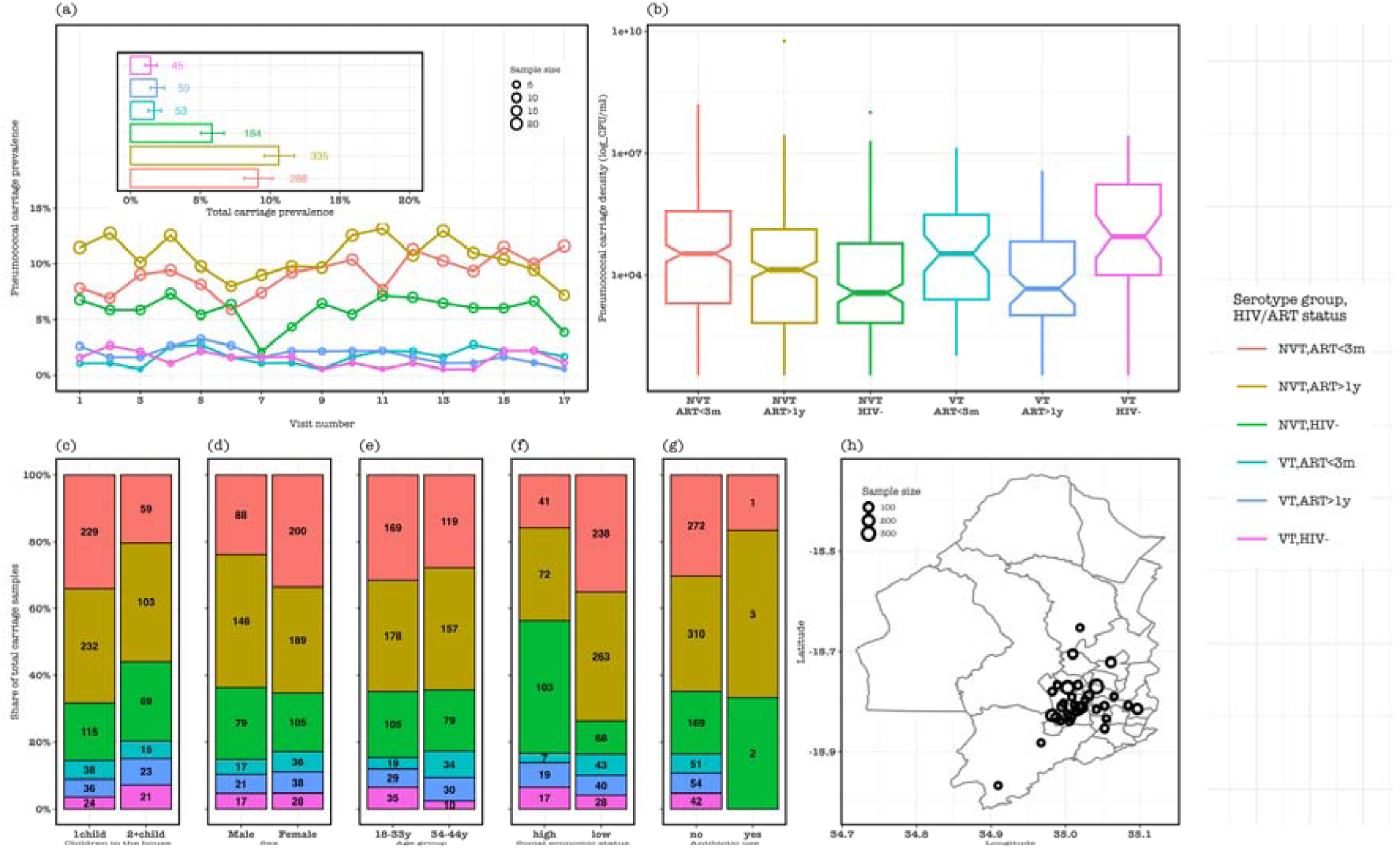
Participant demographic and epidemiologic characteristics of follow-up samples stratified by vaccine-serotype (VT) and non-VT (NVT) carriage and potential risk factors. (a) The prevalence of pneumococcal carriage in all samples at each sampling visit stratified by serotype group and human immunodeficiency virus (HIV) status, with an insert showing pneumococcal carriage prevalence of samples aggregated across all visits. (b) Distribution of pneumococcal carriage density in HIV-adults, adults living with HIV (ALWHIV) on antiretroviral therapy (ART) at most 3 months and at least 1 year. The share of all pneumococcal carriage stratified by serotype group and HIV/ART status among (c) adults living with 1 child or at least 2 children <5y, (d) males or females, (e) 18-33y or 34-44y, (f) low or high social economic status and (g) antibiotic use. (h) Location where nasopharyngeal samples were collected during the study within Blantyre.

### Pneumococcal carriage acquisition dynamics

The probability that NVT carriage would next be acquired in a non-carrier was generally higher than VT carriage (82.9%, 95% CI: 78.3-86.7% vs 17.1%, 95%CI: 13.4-21.7%). Thus, among ALWHIV on ART>1y, ALWHIV on ART<3m and HIV-, we respectively estimated the annual acquisition episodes of 5.9 (95% CI: 4.3-7.9), 6.0 (95% CI: 4.4-7.9) and 5.0 (95% CI: 3.8-6.8) for overall carriage, 6.2 (95% CI: 4.4-8.2), 6.0 (95% CI: 4.1-8.4) and 5.0 (95% CI: 3.5-6.9) for NVT carriage, and 0.69 (95% CI: 0.31-1.57), 0.42 (95% CI: 0.19-1.01) and 0.78 (95% CI: 0.40-1.70) for VT carriage.

In a multivariate analysis, the pneumococcal acquisition rate was higher among females vs males of overall carriage (Hazard Ratio [HR]: 1.64, 95% CI: 1.262-2.12), NVT (HR: 1.53, 95% CI: 1.17-2.01) and VT (HR: 1.96, 95% CI: 1.11-3.49), among low vs high SES of overall carriage (HR: 1.47, 95% CI: 1.12-1.94), NVT (HR: 1.38, 95% CI: 1.03-1.83) and VT (HR: 2.06, 95% CI: 1.13-3.77), among adult living with 2+ vs 1 child <5y of VT (HR: 1.78, 95% CI: 1.05-3.01), and among ALWHIV on ART>1y than HIV-of NVT (HR: 1.43, 95% CI: 1.01-2.02). (Fig 2, Table 2, Table S1).

**Figure 2.**
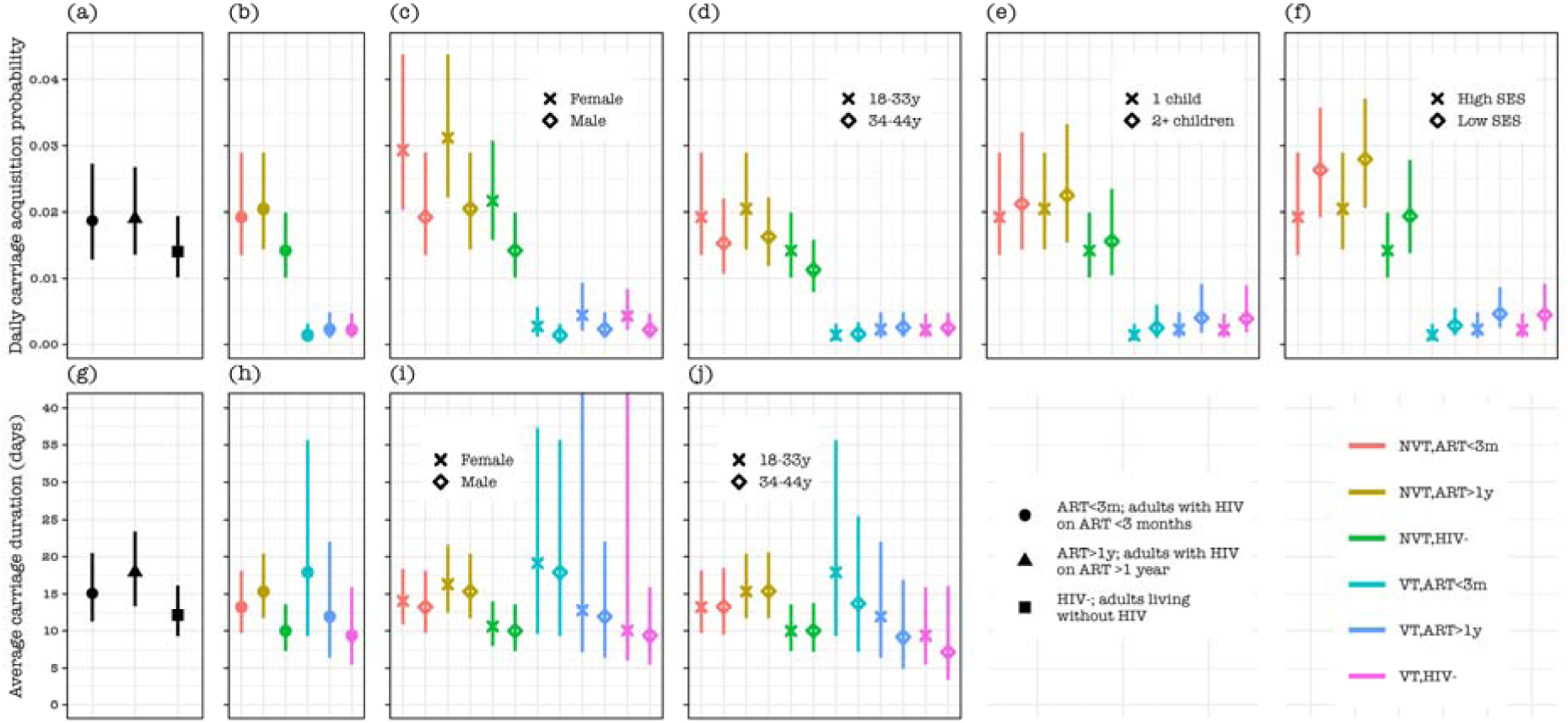
Pneumococcal carriage acquisition probability and duration by serotype group and human immunodeficiency virus (HIV) infection status among potential risk groups. Daily pneumococcal carriage acquisition probability for (a) overall carriage, and carriage stratified by vaccine-serotype group and HIV status among (b) all participants, (c) females or males, (d) adults aged 18-33 or 34-44 years-old (y), (e) adults living with 1 child or at least 2 children in the house, and (f) adults in low or high social economic status (SES). Pneumococcal carriage duration in days for (g) overall carriage, and carriage stratified by vaccine-serotype group and HIV status among (h) all participants, (i) females or males, and (j) adults aged 18-33 or 34-44 years-old (y).

**Table 2.**
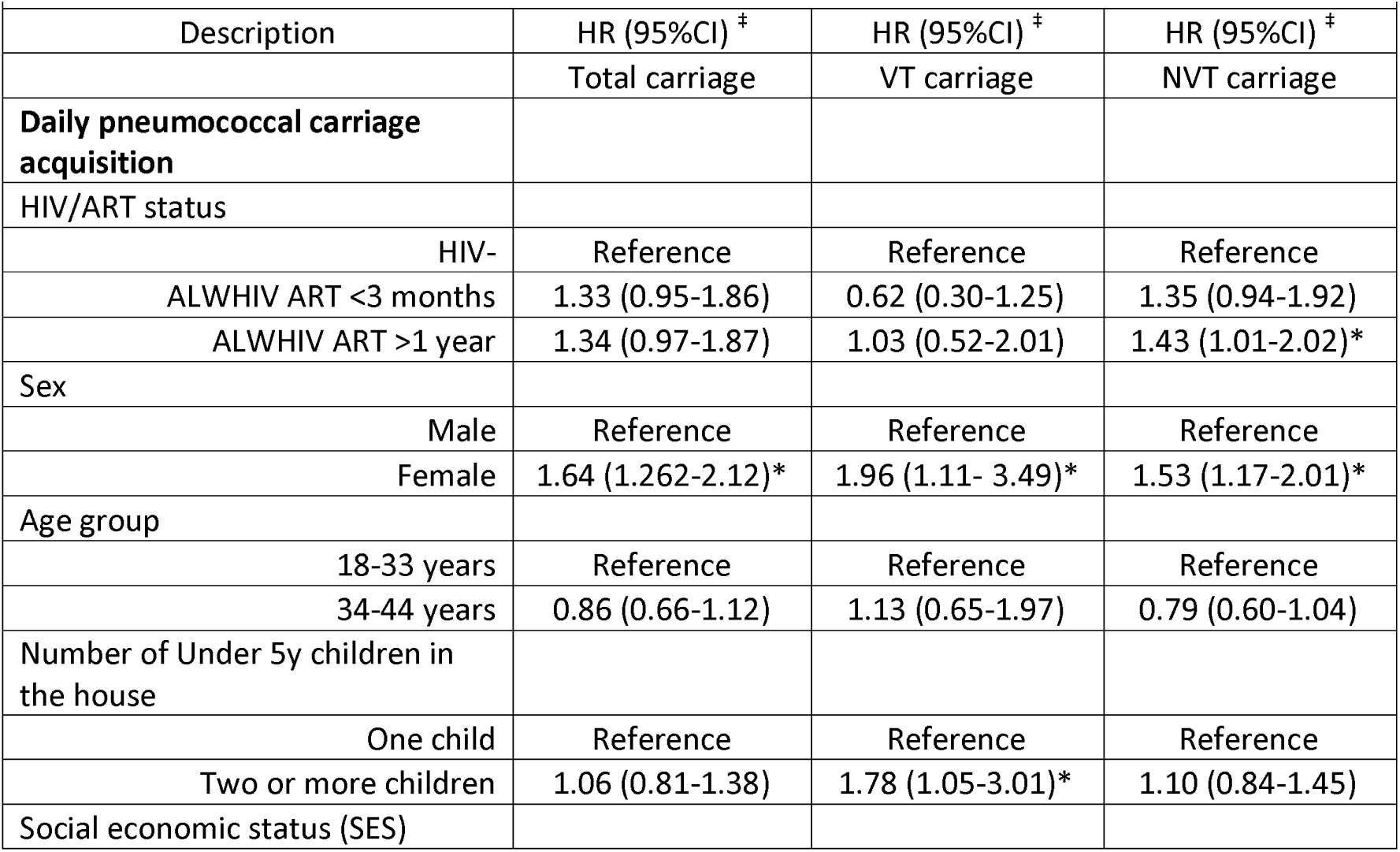
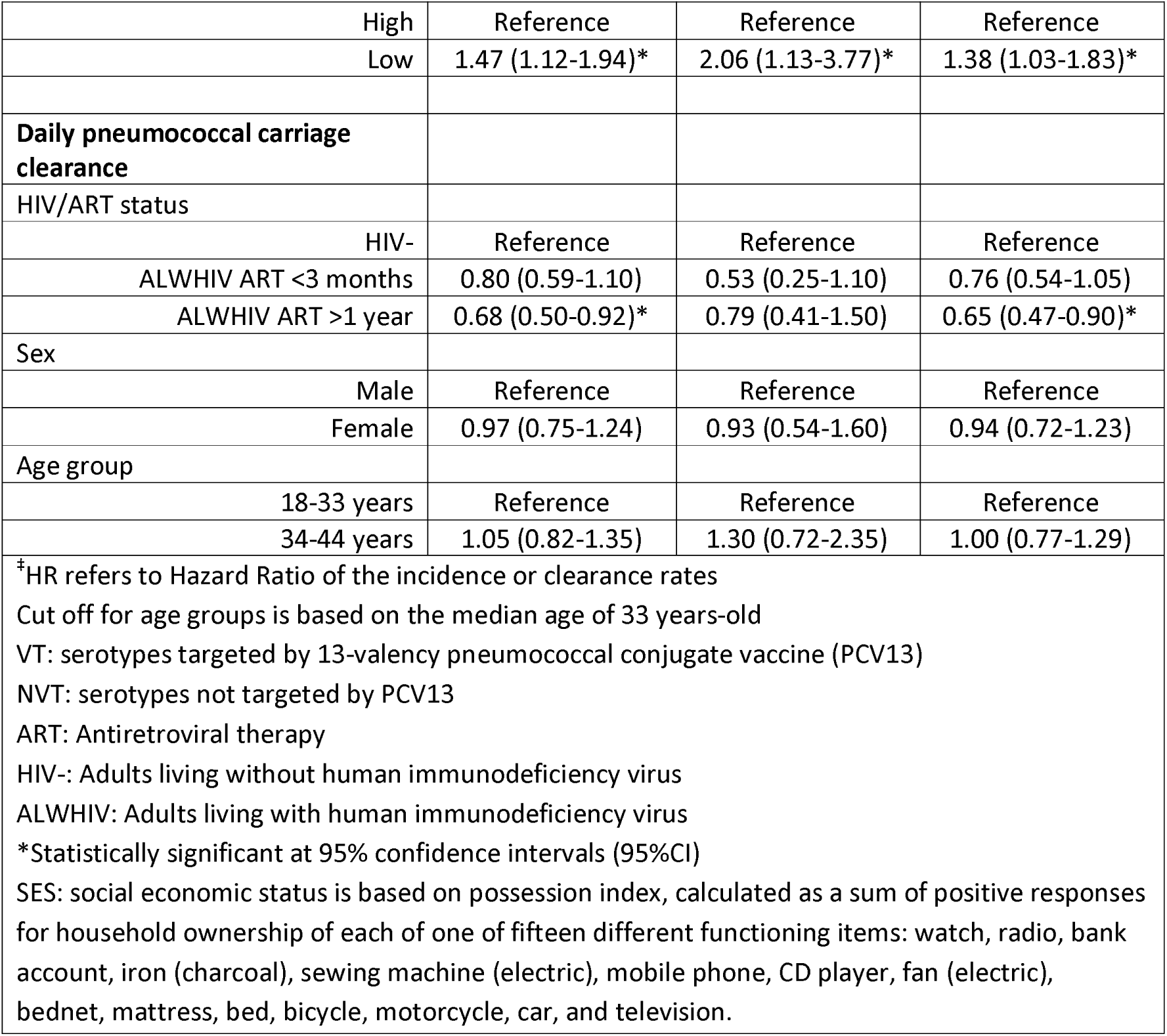
The effect of each considered risk factor on pneumococcal acquisition and clearance rates estimated from the Markov model using data from a longitudinal nasopharyngeal swabbing study conducted in Blantyre, Malawi between September 2021 to December 2023.

### Pneumococcal carriage duration dynamics

The average overall carriage duration was slightly higher among ALWHIV on ART>1y (17.9 days, 95% CI: 13.7-23.6) compared to ALWHIV on ART<3m (15.2 days, 95% CI: 11.2-20.4) or HIV-adults (12.2 days, 95% CI: 9.2-16.1). In a stratified analysis, the average carriage duration was comparable between VT (9.4 days, 95% CI: 5.5-16-0) and NVT (9.9 days, 95% CI: 7.3-13.6) HIV-carriers. However, it was lower for NVT (13.2 days, 95% CI: 9.7-18.1) than VT (17.9 days, 95% CI: 9.3-35.7) in ALWHIV on ART<3m carriers, and higher for NVT (15.4 days, 95% CI: 11.7-20.4) than VT (11.9 days, 95% CI: 6.4-22.0) ALWHIV on ART>1y carriers.

In a multivariate analysis, pneumococcal carriage clearance was slower among ALWHIV on ART>1y compared to HIV-adults for overall carriage (Hazard Ratio [HR]: 0.68, 95% CI: 0.50-0.92) and NVT carriage (HR: 0.65, 95% CI: 0.47-0.90), and comparable between ALWHIV on ART <3 months and HIV-adults for overall carriage (0.80, 95%CI: 0.59-1.10) and NVT (0.76, 95%CI: 0.54-1.05) (Fig 2, Table 2, Table S2).

### Pneumococcal serotype-specific carriage dynamics

In all adults, the sampling frequency of identified pneumococcal serotypes ranged from n=1 (0.01%) for serotype 14 to n=55 (5.7%) for serogroup 15 or serotype 7A/B/C, with n=410 (42.5%) of NVT with unknown serotype (uNVT) e.g., NVT not assigned a specific serotype, being the largest samples. The overall serotype carriage dynamics without stratifying by HIV status showed that serotypes 3 (0.14%, 95% CI: 0.09-0.23) and 19F (0.16%, 95% CI: 0.10-0.26) among PCV13 serotypes, and serogroup 15 (0.18%, 95%CI: 0.12-0.27), serogroup 11 (0.18%, 95% CI: 0.12-0.28) and 23A/B (0.17%, 95%CI: 0.10-0.26) among non-PCV13 serotypes generally had high daily acquisition probability compared to other serotypes or serogroups. On the other hand, serotypes 19A (17.4 days, 95% CI: 7.3-43.1), 3 (13.3 days, 95% CI: 8.6-20.5) and 6A (12.9 days, 95% CI: 5.3-30.5) among PCV13 serotypes, and 7A/B/C (17.8 days, 95% CI: 11.4-28.9), serogroup 15 (15.8 days, 95% CI: 9.6-25.7), and 17A/F (15.0 days, 95% CI: 8.7-26.8) among non-PCV13 serotype were carried the longest. Co-colonisation of pneumococcal serotypes or serogroup (colonisation chains) were more frequent among NVT (e.g., between NVT with known serotypes [kNVT] and uNVT) than between VT and NVT, with the highest colonisation chains estimated between uNVT and serogroup 15 or kNVT (Fig 3, Table S3).

**Figure 3.**
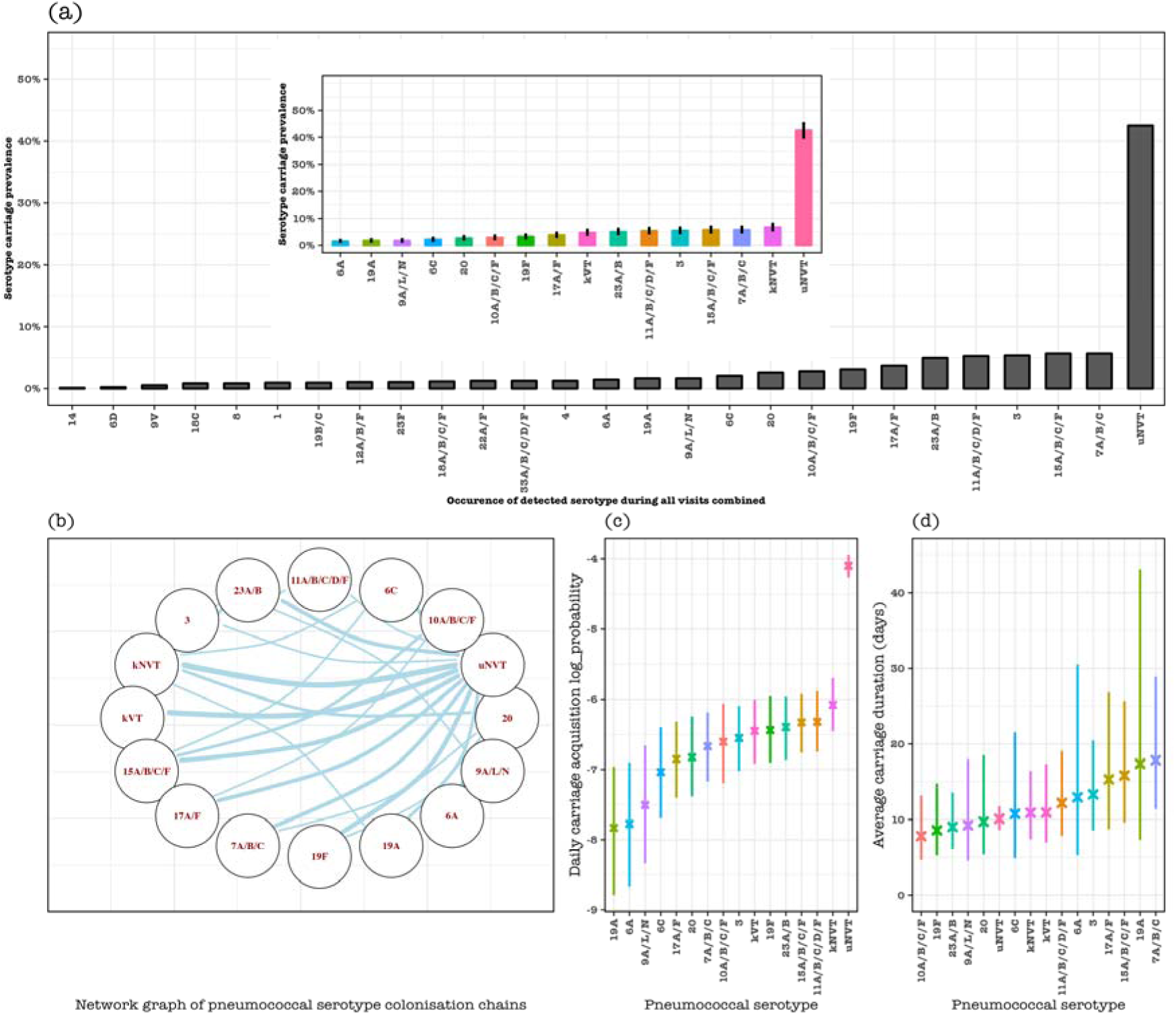
Pneumococcal serotype-specific carriage dynamics in considered serotypes with relative high sampling frequency. (a) Prevalence of each serotype in all samples, with ‘uNVT’ representing unknown non-PCV13 serotypes because they were not included in the serotyping assay which could only identify up to 23 serotypes including all VT. Insert in (a) is the carriage prevalence of each serotype or serogroup with a relatively high sample frequency, where ‘kVT’ represents known PCV13 serotypes with very low sample frequency (1, 4, 9V, 14, 18C and 23F), and ‘kNVT’ represents non-PCV13 serotypes with known serotypes with very low sample frequency (22A/F, serogroup 33 and 18, 12A/B/F, 19B/C, 8 and 6D). (b) Network diagram showing the acquisition of a serotype replacing a specific serotype in a colonisation chain. The size of the edges reflects the pairs of serotype transition events in the colonisation chain that occur more likely than expected, and the node represents the serotype or serotype group or vaccine-serotype group. (c) Daily pneumococcal carriage acquisition probability under a log scale and (d) daily average pneumococcal serotype carriage duration.

## Discussion

We have used multi-state Markov models to disentangle pneumococcal serotype carriage dynamics in ALWHIV and HIV-adults in a mature infant PCV13 program in Malawi. We estimate substantial acquisitions of VT and NVT carriage in females and those living under low socioeconomic status. High VT acquisitions among adults living with at least two children <5Y in the house and NVT acquisitions among ALWHIV on ART>1y are also estimated. On the other hand, prolonged durations of NVT carriage are estimated among ALWHIV on ART>1y. Residual PCV13 serotypes 19F and 3 are highly acquired, whereas 19A, 3, and 6A are prolongedly carried, although non-PCV13 serotypes remain dominant in circulation among adults. Our findings unravel pneumococcal carriage dynamics among ALWHIV and provide baseline estimates for assessing future pneumococcal vaccine impact in ALWHIV. These results suggest that a PCV strategy in ALWHIV with expanded serotype coverage may be warranted to tackle the remaining preventable burden of pneumococcal carriage and subsequent disease in ALWHIV.

Pneumococcal carriage prevalence has previously been reported to be higher among ALWHIV on ART compared to those not on ART in rural Malawi [8]. Our study shows a similar higher prevalence of pneumococcal carriage in ALWHIV on ART>1y than those on ART<3m or HIV-adults. Follow-up studies are required to investigate the biological factors for the increased pneumococcal prevalence in individuals who have been on ART for an extended period compared to those who recently started treatment. We further show that this elevated carriage in ALWHIV on ART>1y is likely influenced by frequent acquisitions and prolonged carriage duration of NVT serotypes.

Antibiotic use is sometimes associated with individual carriage reduction [29], but it’s role was not assessed in this study due to limited data points on antibiotic uptake. Nonetheless, the baseline and follow-up density of pneumococcal carriage remained comparable between HIV groups. Thus, it remains unclear whether the slow NVT clearance is linked to reported cotrimoxazole or penicillin-resistant pneumococci among ALWHIV in this setting [21]. If indeed the reported drugs select for resistant NVT, it may suggest that colonisation of resistant NVT pneumococci in ALWHIV may be inefficiently cleared at the mucosal level, leading to prolonged duration of pneumococcal carriage. However, causal links of prolonged pneumococcal carriage among ALWHIV need further investigation from laboratory measures.

Children <5y remain the major reservoir of pneumococcal carriage transmission in the era of PCV13 in this setting and elsewhere [14, 16, 26, 30]. Since female adults are more likely to interact with younger children due to cultural and parental roles, social mixing is highly intensive between children and females compared to male adults in this setting [31], likely resulting in higher carriage acquisition risk in females than males consistent with our findings in this study. Furthermore, household spread of pneumococcus is usually influenced by higher household density [26, 32, 33], and having more younger children in the house who are a major reservoir of pneumococcal carriage transmission increases the risk of pneumococcal carriage acquisition [3]. Similarly, higher pneumococcal carriage acquisitions in low than high SES households, as shown here and in previous studies in this setting [3], is likely related to poor living conditions, including poor ventilation and overcrowding. However, fine-scale household pneumococcal carriage dynamics, including quantifying the contribution of different household members to pneumococcal carriage transmission, remain a gap to be addressed in this setting.

Serotype-specific pneumococcal carriage acquisition and clearance estimates reported in our study have implications for the choice of PCV strategy in ALWHIV in this setting. PCV13 serotypes still in circulation underscore inadequate herd immunity from the infant PCV13 program [14, 16], and suggest that ALWHIV remains at high risk of preventable pneumococcal carriage and subsequent disease [17]. Thus, providing direct PCV protection to ALWHIV or indirect protection by switching to a new infant PCV schedule that substantially improves herd immunity among ALWHIV is urgently needed [18]. The high presence of NVT implies that ALWHIV have the additional risk of pneumococcal disease that may not be prevented by PCV13, necessitating the need for assessing the impact of a newer infant or ALWHIV PCV products with expanded serotype coverage. Of note, serotypes 1 and 5 cause most pneumococcal invasive disease in children in this setting [34], yet were not isolated in adults in this study reflecting that serotypes circulating in carriage do not usually match those in disease as reported by others[35]. Moreover, the extent to which serotypes circulation in adults influence those in children and vice versa remains to be quantified. Thus, the choice of a PCV strategy partly needs to account for the complex interaction between at risk age groups, PCV serotype coverage, and the distribution in serotype carriage and disease in this setting [36].

Our study did not explicitly account for simultaneous carriage of multiple serotypes because latex agglutination was used for serotyping a single bacterial colony [21]. Absence of multiple serotype detection may have biased downward on acquisition rates by missing acquisition events of new serotype while detecting resident serotype and carriage duration by failing to detect serotype when another dominant serotype is present [2]. Another limitation of this study is the lack of serotyping data for all the NVT serotypes. Follow-up studies should use molecular assays or whole-genome sequencing approaches to reliably detect the carriage of multiple serotypes within an individual [37]. Insufficient data points propelled us to combine the carriage of some serotypes targeted or not targeted by PCV13 to estimate serotype dynamics. The distribution of serotypes in healthy carriers is needed to evaluate PCV impact on invasive disease [19], and our study provides baseline estimates of serotype distribution, acquisition, and clearance at vaccine-serotype group and serotype-specific levels, for assessing future PCV impact in ALWHIV.

In conclusion, the disproportionately high pneumococcal carriage prevalence in ALWHIV on ART>1y is mostly due to high acquisition and prolonged duration of NVT. Our study provides baseline estimates of pneumococcal serotype dynamics for comparison when new PCV strategies are implemented directly in ALWHIV or indirectly in infants.

## Acknowledgements

The authors thank all community study participants, and the study staff for their support and co-operation during the study. This work was supported by an African Research Leader (ARL) award [MR/T008822/1 to KCJ. This ARL award is jointly funded by the UK Medical Research Council (MRC) and the UK Foreign Commonwealth and Development Office (FCDO) under the MRC/FCDO Concordat agreement and is also part of the EDCTP2 programme supported by the European Union. A Wellcome Strategic award number 206545/Z/17/Z supports MLW. The funders were not involved in the design of the study; in the collection, analysis, and interpretation of the data; and in writing the manuscript. The findings and conclusions in this report are those of the authors and do not necessarily represent the official position of the funders.

## Author contributions

Conceptualization; JP, LS, DT, KCJ

Data curation; LS, LM, DT

Formal analysis; DT

Funding acquisition; KCJ

Investigation; LS, LM, DT, KCJ

Methodology; JP, CG, LS, DT, KCJ

Project administration; JP, KCJ

Resources; KCJ

Software; DT

Supervision; LS, JP, KCJ

Validation; KCJ

Visualization; DT

Writing - original draft; JP, LS, DT, KCJ

Writing - review & editing; JP, LS, LM, NM, AK, MK, TK, EL, PC, KM, CC, DMF, DT, KCJ

All authors read and approved the final manuscript.

## Data availability

An R script that was used to analyse the datasets is available in the GitHub repository https://github.com/deusthindwa/markov.model.pneumococcus.hiv.malawi

